# The systemic inflammatory response and clinicopathological characteristics in patients admitted to hospital with COVID-19 infection: Comparison of 2 consecutive cohorts

**DOI:** 10.1101/2021.02.04.21250932

**Authors:** Donogh Maguire, Conor Richards, Marylynne Woods, Ross Dolan, Jesse Wilson Veitch, Wei MJ Sim, Olivia EH Kemmett, David C Milton, Sophie LW Randall, Ly D Bui, Nicola Goldmann, Amy Brown, Eilidh Gillen, Allan Cameron, Barry Laird, Dinesh Talwar, Ian M Godber, John Wadsworth, Anthony Catchpole, Alan Davidson, Donald C McMillan

## Abstract

**Background:** In order to manage the COVID-19 systemic inflammatory response, it is important to identify clinicopathological characteristics across multiple cohorts.

**Methods:** Electronic patient records for 2 consecutive cohorts of patients admitted to two urban teaching hospitals with COVID-19 during two 7-week periods of the COVID-19 pandemic in Glasgow, U.K. (cohort 1: 17^th^ March 2020 – 1^st^ May 2020) and (cohort 2: 18^th^ May 2020 – 6^th^ July 2020) were examined for routine clinical, laboratory and clinical outcome data.

**Results:** Compared with cohort 1, cohort 2 were older (p<0.001), more likely to be female (p<0.05) and have less independent living circumstances (p<0.001). More patients in cohort 2 were PCR positive, CXR negative (both p<0.001) and had low serum albumin concentrations (p<0.001). 30-day mortality was similar between both cohorts (23% and 22%). Over the 2 cohorts, age ≥70 (p<0.001), male gender (p<0.05), hypertension (p<0.01), heart failure (p<0.05), cognitive impairment (p<0.001), frailty (p<0.001), COPD (p<0.05), delirium (p<0.001), elevated perioperative Glasgow Prognostic Score (p≤0.001), elevated neutrophil-lymphocyte ratio (p<0.001), low haematocrit (p<0.01), elevated urea (p<0.001), creatinine (p<0.001), glucose (p<0.05) and lactate (p<0.01); and the 4C score were associated with 30-day mortality. When compared with the 4C score, greater frailty (OR 10.2, 95% C.I. 3.4 – 30.6, p<0.01) and low albumin (OR 5.6, 95% C.I. 2.0 – 15.6, p<0.01) were strongly independently associated with 30-day mortality.

**Conclusion:** In addition to the 4C mortality score, frailty score and a low albumin were strongly independently associated with 30-day mortality in two consecutive cohorts of patients admitted to hospital with COVID-19.

**Article summary:**

- In two consecutive cohorts of patients with COVID-19 infection admitted to two urban teaching hospitals in Glasgow, UK, there were variations in a number of clinicopathological characteristics despite similar mortality (23 and 22%).
- In these two cohorts, in a multivariate analysis that included the 4C mortality score, clinical frailty score >3, low serum albumin concentration (<35 g/L), high neutrophil-lymphocyte ratio (≥5), and abnormal serum sodium concentration (<133/>145 mmol/L) remained independently associated with 30-day mortality.

## Background

The number of people worldwide who are known to have been infected with COVID-19 (SARS-CoV-2 infection) increased from 30 million to 84 million in a twelve week period between September 2020 and January 2021, and the number who have died has almost doubled (1 million to 1.8 million) (1). The severity of this viral disease for an individual is associated with a widespread perturbation of immune, physiological and metabolic parameters (2, 3). These whole-body changes are characteristic of a systemic inflammatory response to tissue injury. Indeed, measures of this systemic inflammatory response have been shown to have prognostic value (4–6). In particular, the 4C mortality score was developed in more than 55,000 patients with COVID-19 and measured the systemic inflammatory response using C-reactive protein (6). Other measures of the systemic inflammatory response such as the neutrophil lymphocyte ratio (NLR) have also been shown to have prognostic value (7). Moreover, the systemic inflammatory response has been shown to be a useful therapeutic target in patients with COVID-19 (8–10). However, to date, as there have been variations in the assessment of the systemic inflammatory response, other important factors may remain to be identified. Experience in consecutive cohorts also remains limited.

The aim of the present study was to compare the 4C mortality score, other measures of the systemic inflammatory response and clinicopathological characteristics in two consecutive cohorts of patients on admission with COVID-19.

## Patients and methods

Electronic patient records for patients who were admitted to two large city teaching hospitals (Glasgow Royal Infirmary (GRI) and the Queen Elizabeth University Hospital (QEUH), Glasgow, U.K.), for two consecutive cohorts, cohort 1 (n=243, 1/4/2020-18/5/2020) (5) and cohort 2 (n=261, 18/5/2020-6/7/2020) were examined for routine clinical, laboratory and clinical outcome data. These teaching hospitals serve urban populations with a high burden of socio-economic deprivation and offer the full spectrum of adult acute receiving specialties to patients over 16 years old. In line with NHS policy, the NHS Greater Glasgow and Clyde Caldicott guardian approved this study. The study protocol (GN20AE307) was approved by the North West England – Preston research ethics committee (20/NW/0336) and registered with clinicaltrials.gov (NCT04484545).

Details of the recruitment of patients between 1/4/2020-18/5/2020 for cohort 1 have been previously described (5). In patients admitted to hospital between 18/5/2020-6/7/2020, age, sex, BMI and polymerase chain reaction (PCR) confirmed evidence of COVID-19 infection at time of discharge or death certification were considered minimal criteria for inclusion in cohort 2.

As per routine clinical practice in the Emergency Department (ED) and Acute Assessment Unit (AAU) in both hospitals, patients were scored on the National Early Warning Score (NEWS) at presentation to triage. NEWS is a validated score of severity of physiological derangement that allocates a score (0–3) to six clinical parameters (pulse rate, blood pressure, respiratory rate, oxygen saturations, requirement for supplemental oxygen and level of responsiveness (alert (A), responding to verbal (V), painful (P) stimuli and unresponsive (U)

AVPU scale)) (11). NEWS determines the triage category and level of immediate treatment that is required at the time of presentation, and the interval to re-administering the NEWS scoring tool according to the score achieved (i.e. the severity of physiological derangement). NEWS >4 and >7 are considered to indicate moderately severe and severe physiological derangement respectively.

The 4C Mortality Score is a validated prognostic score that predicts in-hospital mortality among patients with COVID-19 who are admitted to a general hospital setting in the U.K. (6). It includes eight variables that are readily available at initial hospital assessment: age, sex, number of comorbidities, respiratory rate, peripheral oxygen saturation, level of consciousness, urea level, and C-reactive protein (score range 0-21 points) (see Table 1).

**Table 1.**
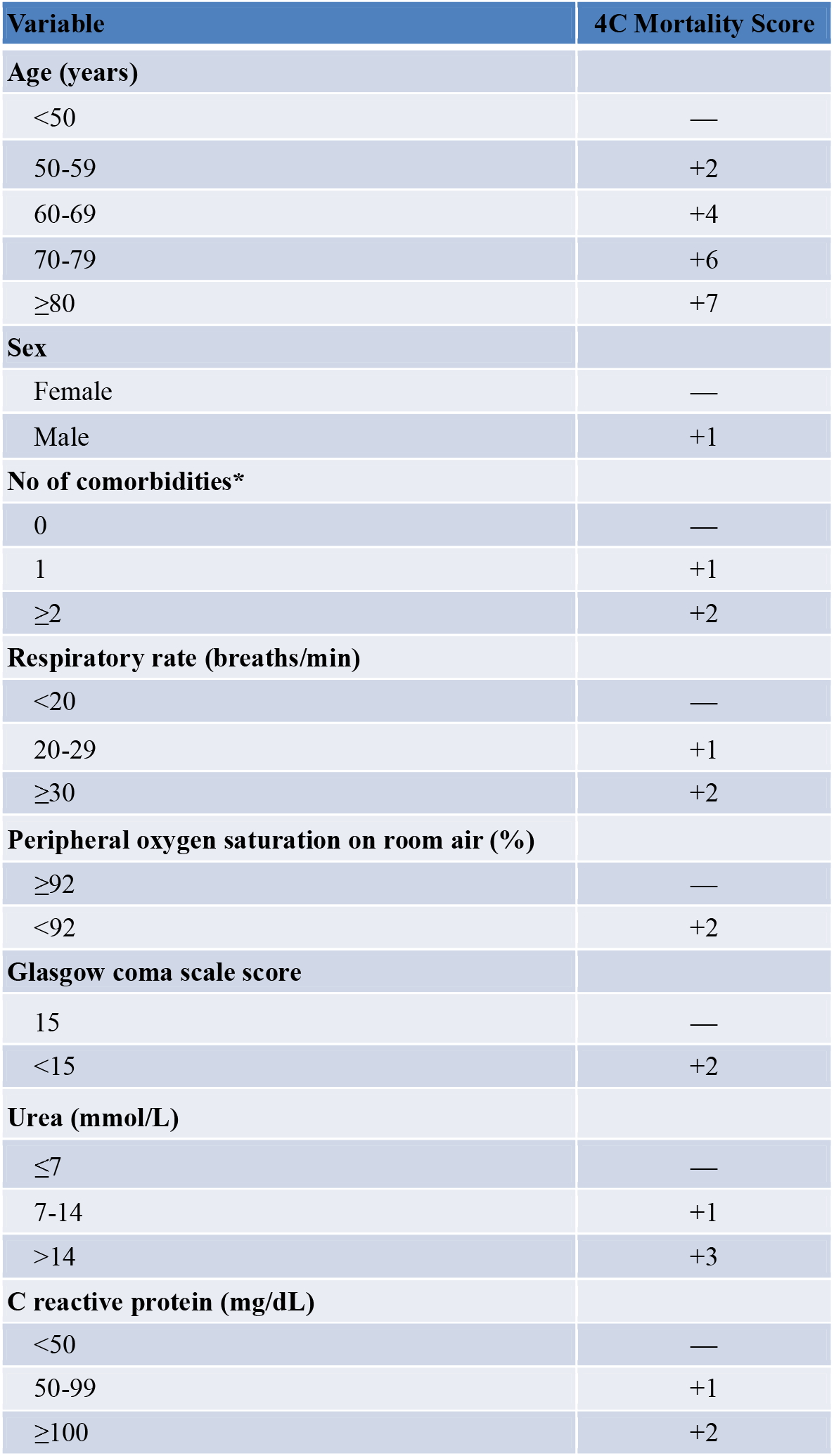
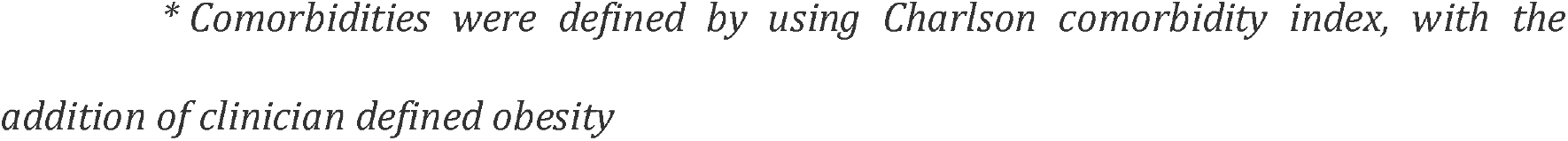
Final 4C Mortality Score for in-hospital mortality in patients with covid-19. Prognostic index derived from penalised logistic regression (LASSO) model (6)

| Variable | 4C Mortality Score |
| --- | --- |
| **Age (years)** |  |
| <50 | — |
| 50-59 | +2 |
| 60-69 | +4 |
| 70-79 | +6 |
| ≥80 | +7 |
| **Sex** |  |
| Female | — |
| Male | +1 |
| **No of comorbidities\*** |  |
| 0 | — |
| 1 | +1 |
| ≥2 | +2 |
| **Respiratory rate (breaths/min)** |  |
| <20 | — |
| 20-29 | +1 |
| ≥30 | +2 |
| **Peripheral oxygen saturation on room air (%)** |  |
| ≥92 | — |
| <92 | +2 |
| **Glasgow coma scale score** |  |
| 15 | — |
| <15 | +2 |
| **Urea (mmol/L)** |  |
| ≤7 | — |
| 7-14 | +1 |
| >14 | +3 |
| **C reactive protein (mg/dL)** |  |
| <50 | — |
| 50-99 | +1 |
| ≥100 | +2 |

In the present study, age was grouped as less than 40 years, 40-49 years, 50-59 years, 60-69 years, 70-79 years and 80 years and older. Age categories were further simplified to </≥ 70 years. Social deprivation was defined by the Scottish Indices of Multiple Deprivation 2019 based on individual home postcode. Ethnicity was classified as White, Mixed, Asian, Black, or other ethnic group.

Frailty was assessed using the Clinical Frailty Scale (CFS) (12, 13). The CFS is a validated measure of clinical frailty that has been shown to have prognostic value (13). The CFS includes items such as comorbidity, cognitive impairment and disability while also incorporating functional interpretation of physical frailty according to level of dependence in living circumstances (12). In the present study, living circumstances were classified as: independent; living at home with support from family member / paid carer or sheltered accommodation; care home; or dependent living in a nursing home.

Admission serum C-reactive protein (CRP), albumin concentrations and differential blood cell counts were categorised using local reference intervals. Neutrophil-lymphocyte ratio (NLR) and the peri-operative Glasgow Prognostic Score (poGPS) were calculated as outlined in Tables 2 and 3 (14–16). The NLR and poGPS are validated prognostic scoring systems that have been used in a variety of clinical settings. They both utilise two components, neutrophils/ lymphocytes and C-reactive protein/ albumin respectively, that are routinely measured in patients admitted to the general hospital setting. For this study, each scoring system had 3 divisions indicating mild, moderate and severe systemic inflammatory response respectively (17).

**Table 2.** Calculation of the Neutrophil Lymphocyte Ratio (NLR)

| Neutrophil Lymphocyte Ratio (NLR): | Ratio | SIRS Severity |
| --- | --- | --- |
| Neutrophil count: lymphocyte count | <3 | Mild |
| Neutrophil count: lymphocyte count | 3-5 | Moderate |
| Neutrophil count: lymphocyte count | >5 | Severe |

**Table 3.** Peri-operative Glasgow Prognostic Score (poGPS)

| peri-operative Glasgow Prognostic Score (poGPS) | Score | SIRS Severity |
| --- | --- | --- |
| C-reactive protein ≤ 150mg/l and Albumin ≥25 g/l | 0 | Mild |
| C-reactive protein > 150mg/l and Albumin ≥25 g/l | 1 | Moderate |
| C-reactive protein ≤ 150mg/l and Albumin <25 g/l | 1 | Moderate |
| C-reactive protein > 150mg/l and Albumin <25 g/l | 2 | Severe |

## Statistical Analysis

Demographical and clinicopathological data were presented as categorical variables using recognised clinical thresholds. These variables were analysed using χ^2^ test for linear-by-linear association, or χ2 test for 2-by-2 tables.

Associations between demographical, clinicopathological characteristics and mortality were analysed using univariate and a multivariate backward conditional approach. A *p* < 0.05 was applied to inclusion at each step in the multivariate analysis.

A convenience sampling strategy was adopted based on the patients admitted during the study period; therefore, a formal sample size calculation was not performed. Missing data were excluded from analysis on a variable-by-variable basis. Two-tailed *p* values <0.05 were considered statistically significant. Statistical analysis was performed using SPSS software version 27.0. (SPSS Inc., Chicago, IL, USA).

## Results

Details of the recruitment of patients for cohort 1 (n=243) have been previously described (5). In cohort 2, of the 356 patients who were confirmed to have COVID-19 infection by PCR test, 278 patients fulfilled the criteria for inclusion with age, sex, BMI. Seventeen patients were re-admitted and these were excluded from the analysis at second admission leaving 261 patients to be included in the analysis.

Comparison of the demographical and clinicopathological characteristics of the two cohorts are shown in Table 4. Compared with cohort 1, cohort 2 were older (p<0.001), more likely to be female (p<0.05) and have less independent living circumstances (p<0.001). With reference to previous medical history, compared with cohort 1, cohort 2 had hypertension and heart failure (both p<0.05), had chronic renal failure (p<0.001), had cognitive impairment and previous delirium (both p<0.01), were less frail (p<0.001) and had less asthma (p<0.01). With reference to diagnostic criteria, compared with cohort 1, cohort 2 were more likely to be PCR positive and CXR negative (both p<0.001). With reference to laboratory results, compared with cohort 1, cohort 2 had low albumin (p<0.001), low haemoglobin (p<0.001), low haematocrit (p<0.05), lower MCV (0.05), abnormal sodium (p<0.01), elevated creatinine (p<0.01), elevated alkaline phosphatase (p<0.001). 30-day mortality was similar between the cohorts (23% and 22%).

**Table 4.**
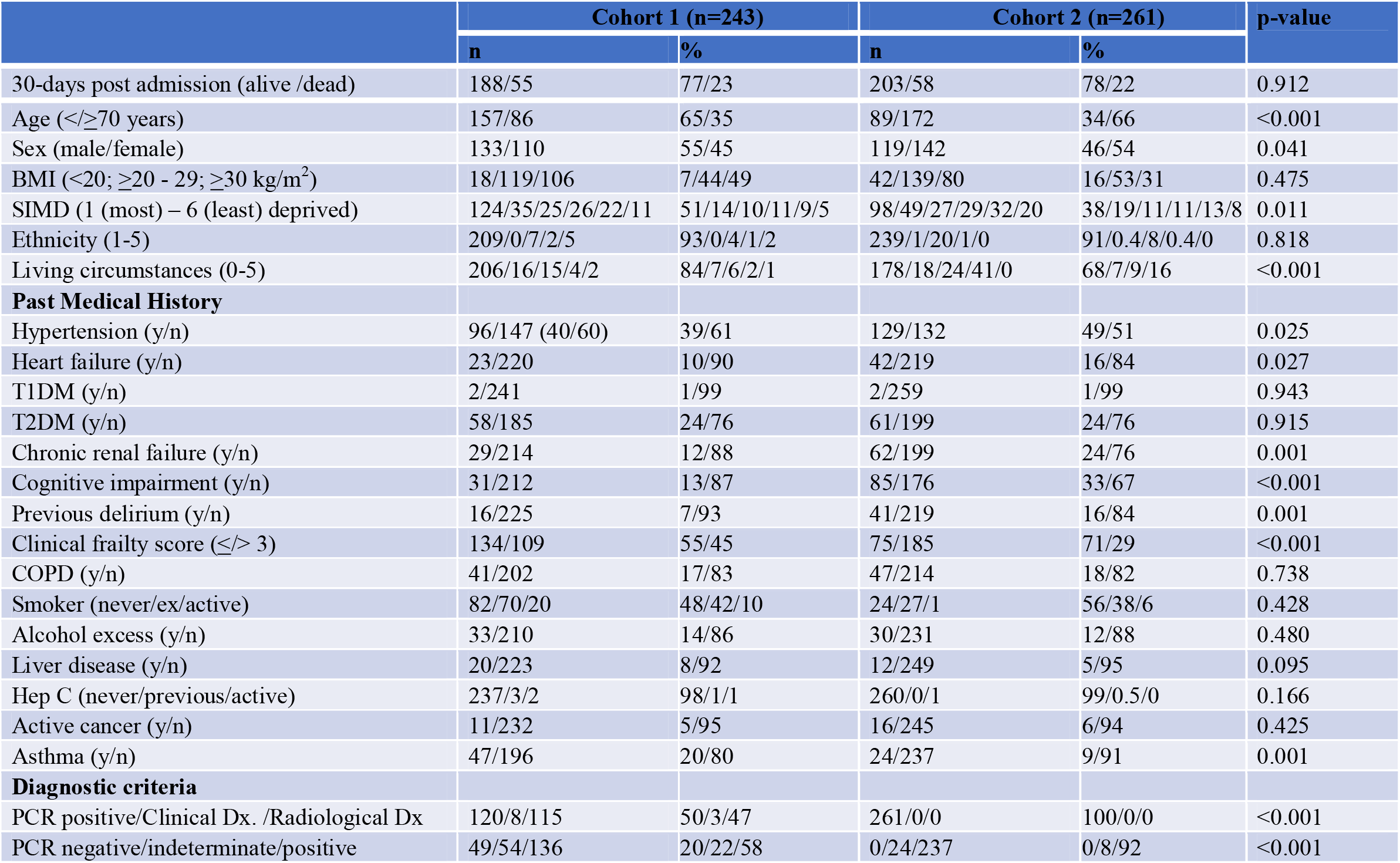
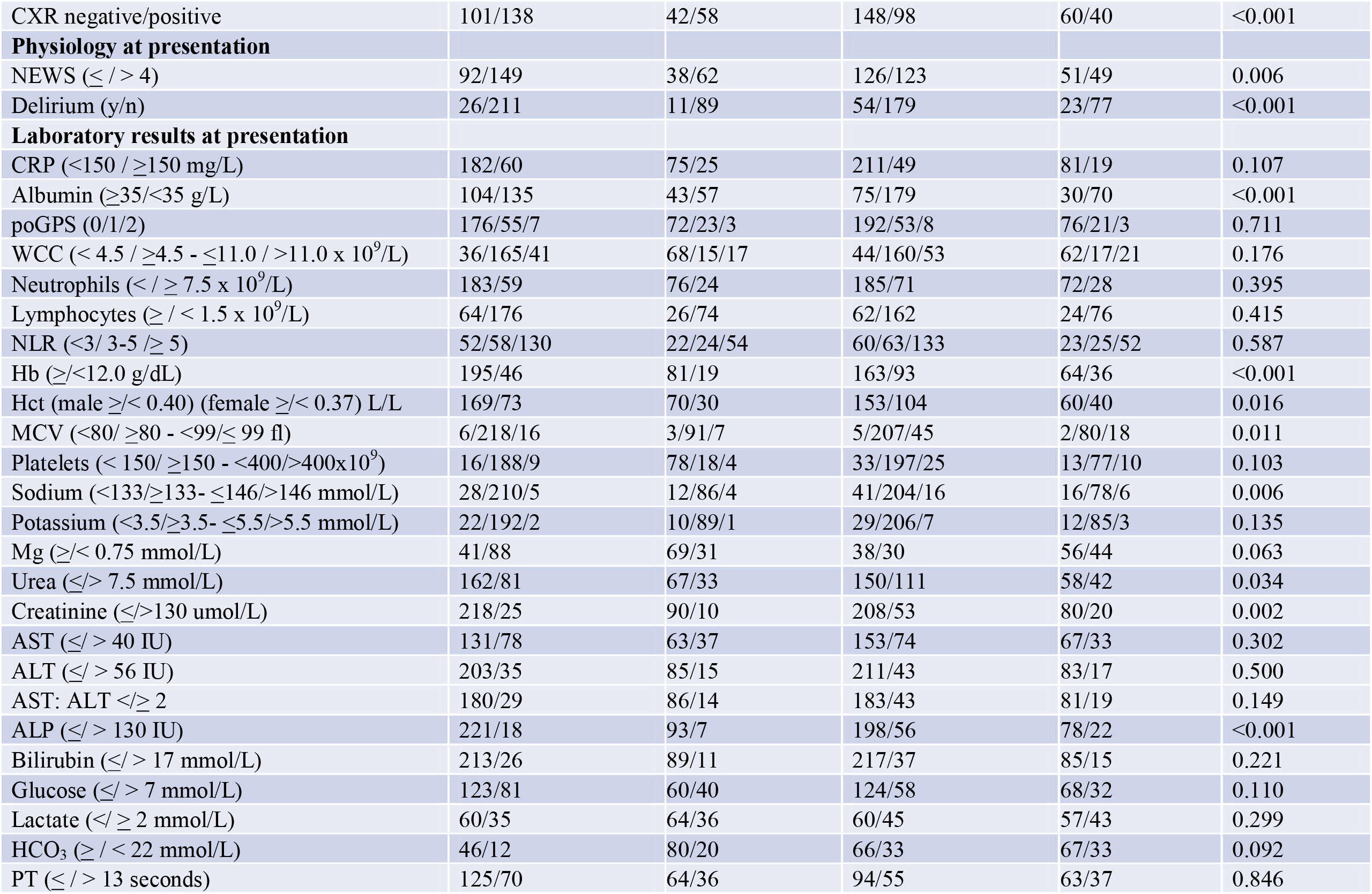
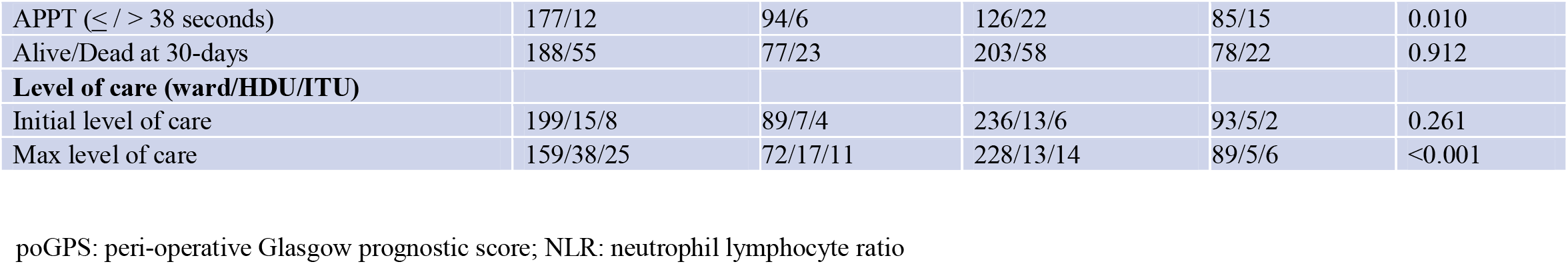
Comparison of the demographical and clinicopathological characteristics of 2 consecutive cohorts patients admitted to hospital with COVID-19.

The relationship between demographic and clinicopathological characteristics and 30-day mortality in the two combined cohorts is shown in Table 5. Over the 2 cohorts, age ≥70 years (p<0.001), males (p<0.05), hypertension (p<0.01), heart failure (p<0.05), cognitive impairment (p<0.001), frailty (p<0.001), COPD (p<0.05), less asthma (p<0.01), delirium (p<0.001), elevated poGPS (p≤0.001), elevated NLR (p<0.001), low haematocrit (p<0.01), abnormal sodium (p<0.001), elevated urea (p<0.001), elevated creatinine (p<0.001), elevated glucose (p<0.05, elevated lactate (p<0.01), elevated PT (p<0.05) and the 4C score were associated with 30-day mortality.

**Table 5.**
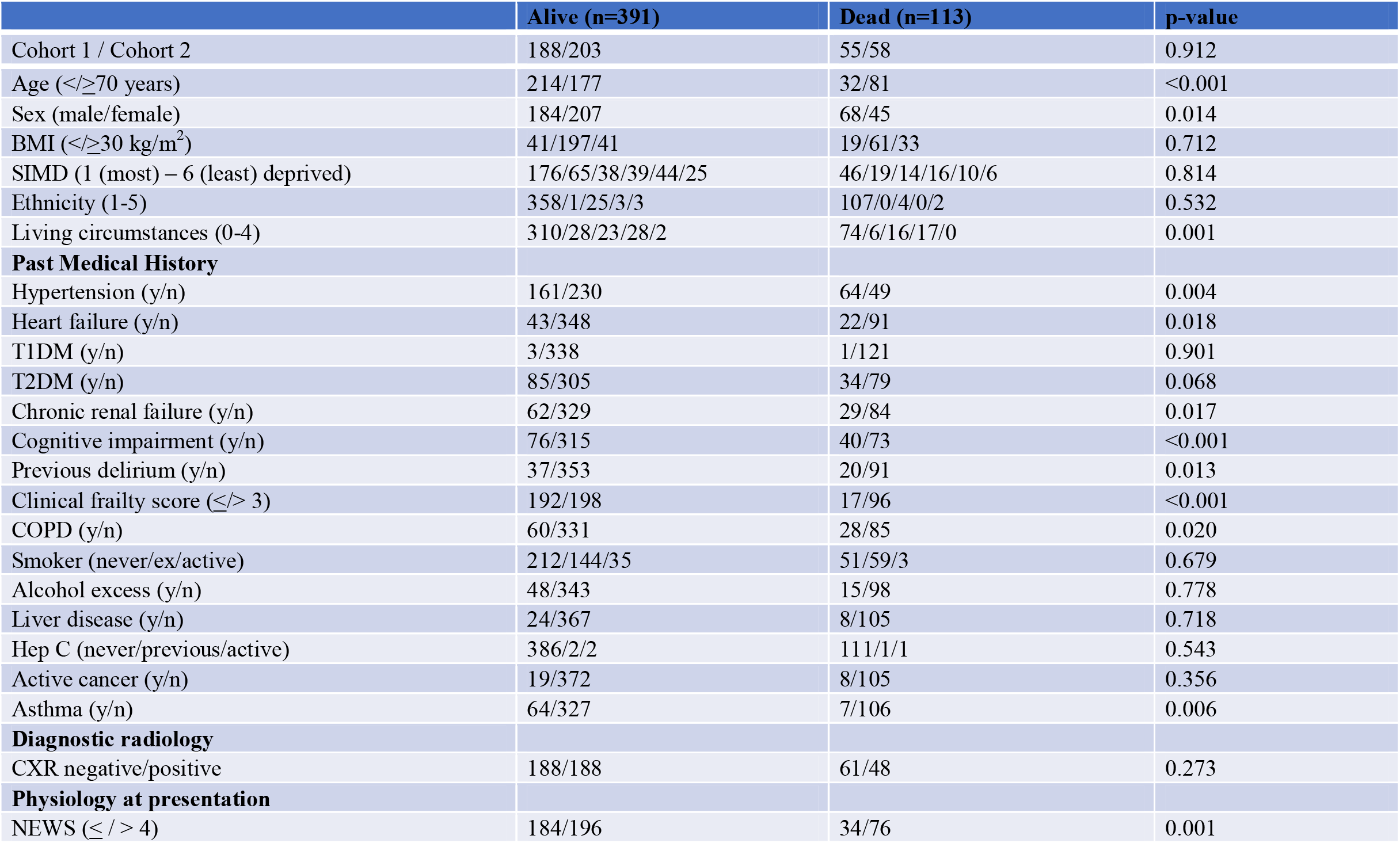
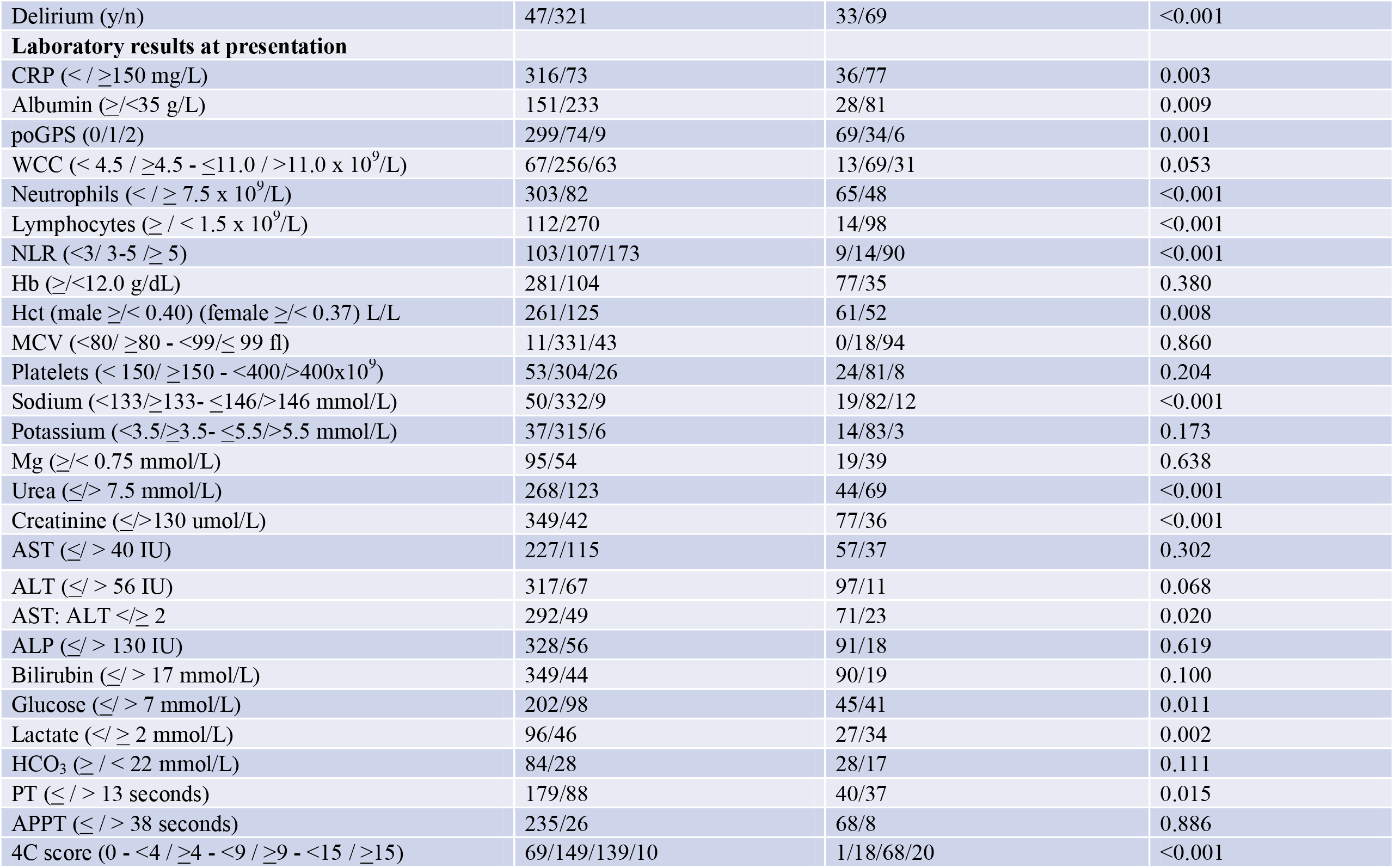
Univariate analysis of demographical, clinicopathological characteristics and 30-day mortality in the two combined cohorts of patients admitted with confirmed COVID-19 (n=504).

To determine which admission parameters were independently associated with 30-day mortality, those factors identified in Table 5 as significant and not in the 4C mortality score were also entered into a binary logistic regression analysis (Table 6). In this multivariate analysis of a restricted dataset (due to limited lactate measurements), only greater frailty (OR 10.2, 95% C.I. 3.4 – 30.6, p<0.01), low albumin (OR 5.6, 95% C.I. 2.0 – 15.6, p<0.01), high NLR (OR 2.2, 95% C.I. 1.1 – 4.6, p<0.05) and abnormal sodium (OR 2.7, 95% C.I. 1.1 – 6.4, p<0.05) remained independently associated with 30-day mortality.

**Table 6.** Binary logistic regression analysis of demographical, clinicopathological characteristics and 30-day mortality in the two combined cohorts of patients admitted with confirmed COVID-19 (n=203).

|  | Alive (n=142) | Dead (n=61) | p-value | .R. | 95% CI | p-value |
| --- | --- | --- | --- | --- | --- | --- |
| Clinical frailty score (≤/$>$ 3) | 79/63 | 12/49 | <0.001 | 10.2 | 3.4 – 30.6 | <0.001 |
| Albumin (≥/$<$30 g/L) | 119/23 | 39/22 | 0.002 | 5.6 | 2.0 – 15.6 | 0.001 |
| NLR ($<$3/$>$3 - $<$5/≥ 5) | 28/37/77 | 3/8/50 | <0.001 | 2.2 | 1.1 – 4.6 | 0.028 |
| Hct (male ≥/$<$ 0.40) (female ≥/$<$ 0.37) L/L | 106/36 | 35/26 | 0.015 | | | 0.915 |
| Sodium ($<$133/≥133- ≤146/$>$146 mmol/L) | 15/123/4 | 12/41/8 | <0.001 | 2.7 | 1.1 – 6.4 | 0.029 |
| Glucose (≤/$>$ 7 mmol/L) | 80/49 | 24/30 | 0.029 | | | 0.847 |
| Lactate ($<$/≥ 2 mmol/L) <sup>b</sup> | 96/46 | 27/34 | 0.002 | | | 0.765 |
| PT (≤/$>$ 13 seconds) | 71/41 | 22/27 | 0.029 | | | 0.599 |
| 4C score (0 - $<$4 / ≥4 - $<$9 / ≥9 - $<$15 / ≥15) | 19/56/50/6 | 0/6/38/14 | <0.001 | | | 0.153 |

Hct Haematocrit; PT prothrombin time

## Discussion

The results of the present study show that, in 2 consecutive cohorts, there was a variation in the admission demographic and clinicopathological characteristics. In particular, the latter cohort were older, had more cardiovascular and renal disease and greater derangement of their laboratory data. Despite this, 30-day mortality was similar between the cohorts. In both cohorts the 4C score, which incorporates age, sex, comorbidity, respiratory, renal and brain function and a measure of the activation of the systemic inflammatory response, had prognostic value. In addition, when compared directly with the 4C mortality score a number of other factors had independent prognostic value, in particular clinical frailty and a low albumin. Taken together, the present results would suggest that the relationship between clinicopathological factors and short-term mortality in patients with COVID-19 may vary with time. Also, that there are other important factors in short term mortality of patients with COVID-19, not captured by the 4C mortality score and such factors may improve the prognostic value of the 4C score. Therefore, there is a need for further work to determine independent prognostic value of clinicopathological factors in patients presenting with COVID-19.

The basis of the strong prognostic value of the clinical frailty scale and low albumin, independent of the 4C mortality score in these patients, is not clear. However, COVID-19 patients with a clinical frailty score >3 may be considered vulnerable and frail and this entity (the ability to care for themselves and its relationship with mortality) may not be captured directly by the 4C score. In particular, the present results may indicate that having COVID-19 induced cytokine storm in a frail patient is a life threatening event (18). Similarly, with reference to a low serum albumin concentration, a cytokine storm would increase the likelihood of mortality. In the case of a low albumin it is clear that this may reflect both an ongoing systemic inflammatory response and also poor nutritional status (19). It may be that the strong prognostic value of frailty also reflects poor nutritional status since a systemic inflammatory response occurring against a background of low metabolic reserves is likely to lead to cellular and organ dysfunction. If this was the case, then it might be expected that frail and hypoalbuminaemic patients would benefit most from treatment with anti-inflammatory agents and nutritional supplementation. Therefore, it may be important to also consider nutritional risk in patients with COVID-19. Irrespective, it would be important to consider frailty and a low albumin in the assessment of patients with COVID-19 (20).

To date, in patients with COVID-19, there has been a great deal of focus on the virus itself. However, it is clear from the prognostic value of host physiology and the host systemic inflammatory response in the 4C work (6) and in the efficacy of dexamethasone treatment (13) that host factors are of considerable importance in outcome of patients with COVID-19. From the present results it is also clear that frailty and nutritional status are important characteristics to be taken into the “staging” of patients presenting with COVID-19.

## Limitations

The present study has a number of limitations. The sample size is relatively small and therefore subject to limitations such as sample bias. However, the clinicopathological data collected was comprehensive across two cohorts, included factors validated in large cohorts of patients with COVID-19 and therefore allowed direct comparison of these factors.

## Conclusion

In the two consecutive cohorts there were variations in a number of clinicopathological characteristics despite similar mortality. In these two cohorts, in addition to the 4C mortality score, NLR>3, abnormal serum sodium concentration (<133 / >146 mmol/L), clinical frailty score >3 and low serum albumin concentration (<30 g/L) were independently associated with 30-day mortality in patients admitted to hospital with COVID-19 infection.

## Data Availability

Data will be available via clinical trials. com. Authors must be cited.

https://clinicaltrials.gov/ct2/show/NCT04484545

## Abbreviations

(SARS-CoV-2 infection): COVID-19
(SIRS): systemic inflammatory response syndrome
(ISARIC): International Severe Acute Respiratory and emerging Infections Consortium
(WHO): World Health Organization
(CCP-UK): Clinical Characterisation Protocol UK
(4C mortality score): study (performed by the ISARIC Coronavirus Clinical Characterisation Consortium—ISARIC-4C)
(NEWS): national early warning score
(PCR): Polymerase chain reaction
(poGPS): peri-operative Glasgow Prognostic Score
(NLR): neutrophil lymphocyte ratio
(CRP): serum C-reactive protein
(GRI): Glasgow Royal Infirmary
(QEUH): Queen Elizabeth University Hospital
(ED): Emergency Department
(AAU): Acute Assessment Unit
(CXR): chest X-ray
(BMI): Body mass index
(CFS): Clinical Frailty Scale
(AVPU scale): Alert (A), responding to verbal (V), painful (P) stimuli and unresponsive (U)
(SIMD): Scottish Indices of Multiple Deprivation
(χ^2^ test): Chi-squared test

## Declaration

In line with NHS policy, collection and analysis of data was approved by the NHS Greater Glasgow and Clyde Caldicott guardian. Ethics committee approval was obtained and requirement for patient consent was waived for this retrospective case note review. All authors have consented to publication and are guarantors of the manuscript and data presented. Anonymised data will be made available on reasonable request to the corresponding author. None of the authors have any conflict of interest to declare. DT is funded by the Scottish Trace Elements and Micronutrients Diagnostic and Reference Laboratory. DMcM is funded by the University of Glasgow.

## Author Contributions

DM, DCM, RD, DT and BL conceived the idea for the study. DM, DCMM, RD, DT, IG, AC, JW, AD and BL contributed to the study design. MW, CR, JWV, WMS, OEK, DCM, SLR, LDB, NG, AB and EG performed manual data extraction from the electronic patient records. AC performed post-code analysis and deprivation scoring. DM performed the statistical analysis.

## Acknowledgements

The research team wish to acknowledge the assistance of Mrs. Jill Dempster (Project Management Unit, Research and Development Department, Greater Glasgow and Clyde) for her expertise and dedication in relation to this work.

